# A QUALITATIVE EXPLORATION OF PATIENT EXPERIENCE OF SPEECH AND LANGUAGE THERAPY FOR DYSARTHRIA IN THE ACUTE PHASE POST STROKE

**DOI:** 10.1101/2025.09.09.25335432

**Authors:** Sasha B. Archer, Joan K-Y. Ma

## Abstract

**Background:** Dysarthria is a neurological speech impairment which can have a pervasive impact on an individual’s independence, psychological well-being and social participation following a stroke. Speech and language therapy (SLT) in the acute hospital phase aims to provide the stroke survivor with the means to communicate effectively and reintegrate socially following their discharge. However, there is limited understanding of how patients perceive their experience of SLT at this time and how effectively it addresses their needs.

**Aim:** To explore the patient’s experience of dysarthria in the acute hospital phase following a stroke and perceptions of the SLT received during this time.

**Methods:** Participants for this qualitative study were recruited as hospital in-patients from the acute SLT service caseload. Semi-structured interview data was analysed with reflexive thematic analysis and themes constructed.

**Results & Discussion:** The themes of speech as a competing priority, feeling disempowered in rehabilitation and the need for human connection and their relevance for clinical practice are discussed. Speech difficulties are perceived as one of several priorities competing for participants’ time and energy with the impact of speech often minimised. Feelings of disempowerment come from both internal and external factors, but the importance of hope and belief in potential recovery was emphasised. The expressed need for human connections highlights the need for positive social communication experiences and the value of a communication affirming rehabilitation setting.

**Conclusions:** These themes highlight the multidimensional impact of dysarthria and how it is inextricably linked to the wider psychological, and social dimensions of stroke. SLT during acute hospital rehabilitation must go beyond the physical impairment and address the patient’s evolving priorities, their self-esteem, capacity for positive self-management and social reintegration.

## INTRODUCTION

### Dysarthria post-stroke

Stroke is one of the leading causes for disability in the UK, with an estimated 152,000 people experiencing strokes each year (Stroke Association, 2018). While emerging interventions such as thrombolysis and thrombectomy are associated with improved recovery of stroke (Ali et al., 2015), the increased life expectancy and number of stroke survivors means that we can expect increasing numbers living with the sequalae of a stroke (Wafa et al., 2020). Approximately 52% of stroke survivors will experience acute dysarthria (Mitchell et al., 2021) with up to 70% reporting dysarthria one year post stroke (Baker et al., 2022). Dysarthria is a neurological motor speech disorder characterized by imprecision, weakness, slow or uncoordinated speech movements. The vital role of speech in self-expression and social participation, means that even an objectively mild dysarthria can have profound social and emotional consequences, highlighting the need for early identification and intervention in the acute hospital. Yet there is a lack of robust clinical evidence to guide the delivery of acute speech and language therapy (SLT) based on what approaches patients find useful.

The acute hospital phase following a stroke combines acute medical care with stroke rehabilitation. This is a challenging time for stroke survivors, as they must come to terms with their medical condition, it’s cognitive and physical impact and the potential for long-term disability. SLT in this acute phase aims to enable patients to adapt to their speech limitations, provide functional support, and facilitate early therapeutic interventions. Early supported discharge may cut short rehabilitation time (Conway & Walshe, 2015) and the pressure on NHS resources may mean SLT input prioritises the more pressing medical need of swallowing difficulties. However early intervention for dysarthria plays a critical role in helping patients regain confidence and maintain a sense of control over their recovery. SLT during the acute phase is important for initial recovery and psychological adjustment, but reduced frequency or amount of SLT, means an effective SLT service must ensure it is responsive and adaptable to individual patient needs (Conway & Walshe, 2015).

### Psychosocial impact of dysarthria

Patient experience of acute dysarthria varies. Some patients report resilience and adapt well, while others report profound impacts on their quality of life (Brady et al., 2011b). Interestingly, the subjective severity of dysarthria does not predict the extent of perceived communicative difficulties (Walshe, 2003; Hartelius et al., 2008) or the psychosocial impact (Atkinson-Clement 2019). People with dysarthria describe far-reaching psychosocial consequences, with their speech difficulties negatively impacting relationships (Walshe & Miller, 2011), social interactions (Brady et al., 2011a) and family dynamics (Miller et al., 2006; Gawulayo et al., 2021). People with dysarthria report stigmatisation and changes to identity (Walshe et al., 2008), parental roles (Dickson et al., 2008; Walshe, 2010), and structural and attitudinal barriers (Walshe & Miller, 2011), including significant difficulties returning to meaningful employment (Garcia et al., 2002; Hommel et al., 2009). Brady et al. (2011b) suggest that the therapy approach should not be based on intelligibility alone, but also on the impact that the dysarthria has on the individual.

### Dysarthria therapy in the acute hospital

Although their usefulness to patient outcomes is uncertain (Sellars et al., 2002), therapeutic interventions such as articulation exercises, respiratory control strategies, and compensatory communication techniques remain prevalent (Collis & Bloch, 2012). These approaches to SLT focus on improved intelligibility, i.e. how understandable the speaker is, and use a range of treatment techniques (Skeat, 2007) to increase the strength and precision of movements required for speech (McCauley et al. 2009), pacing to allow for greater speech articulation and more time for listener processing (MacKenzie & Lowitt, 2012), and increased breath support to support increased volume (Spencer et al., 2003). In contrast, the evidence for nonspeech articulation exercises for improving speech production is equivocal at best (MacKenzie et al., 2014). Indeed they may exacerbate increased muscle tone (Murdoch et al., 2009) or be ineffective due to coordination difficulties (Rosenbek & Jones, 2009). Neuromuscular electronic stimulation may be a promising development, but a lack of randomized controlled trials, longer-term follow-up, and quality of life outcomes (Schreiber 2023), means this hasn’t translated to clinical practise (Miller & Bloch, 2017). Delivered “intensively” (i.e. multiple or longer sessions in a short time-frame) there is evidence for the effectiveness of physiological treatments at improving intelligibility (Mahler & Ramig, 2012; Park et al., 2016), particularly in progressive dysarthria (Mendoza Ramos et al., 2021). However, this is physically and emotionally demanding, and arguably comparable improvements may be achieved with less intensive input (Wenke et al., 2011). Furthermore, intensive SLT is often precluded in the acute phase by co-morbidities (Conway & Walshe, 2015), competing therapy and patient factors, such as low mood or anxiety (Brumfitt, 2009), cognitive changes (British Psychological Society, 2010) and fatigue (Riley et al., 2021). Repetition is the key to stimulating neural adaption (Ludlow, 2008; Schrivener et al., 2012), which is the mechanism the brain uses to rewire and heal itself after stroke. Taking all this in account, there is little justification for therapy which directly targets the speech difficulty but fails to meet the increasing demands required for neural adaptation (Clark, 2003; Rosenbek & Jones, 2009). However, given that therapy is guided by patient centred goals, the prevalence of interventions directly targeting speech may be more to do with patient expectations (Dean & Heron, 2010; MacKenzie et al., 2013) than to the evidence base (Finch et al., 2020).

While there is some quantitative evidence to guide intervention in stroke related dysarthria (Palmer & Enderby 2007), the prevalence of comorbidities such as language impairment (Mitchell, 2021) and swallowing difficulties (Bahia, 2016) as well as reduced compliance (Sellers et al., 2002), insight and motivation (Conway & Walshe, 2015) means quantitative research in the acute phase is challenging. This has resulted in limited guidance for evidence-based practice or for specific treatment techniques in the acute phase. Mitchell et al.’s (2017) systematic review found little evidence of long-term benefits of early SLT intervention for dysarthria on patient outcomes. However, this may reflect the priority put on quantifiable outcome data (Walshe, 2010), neglecting the multifactorial impact of dysarthria beyond the level of impairment and intelligibility (Clarke & Black, 2005). In focussing on longer term outcomes, the early psychological benefits of acute SLT may be missed, particularly in a population whose experience is rarely captured as they are often excluded from research due to communication difficulties (Hilari & Northcout, 2006) or poor participant consent rates (Rosewilliam et al., 2015). This means there is little information on the emotional and social challenges patients face in the days and weeks immediately following their stroke and these are often overlooked in goal setting (Rosewilliam et al., 2015).

Establishing basic communication strategies early on in rehabilitation to compensate for speech difficulties can boost patients’ confidence and help them regain a sense of agency over their interactions with significant others as well as healthcare providers. Reducing negative adaptive strategies and misunderstandings improves motivation (Hole, 2012), improving engagement with rehabilitation generally and supporting social reintegration. Social participation is closely related to quality of life (Walshe et al., 2009) and can be improved through environmental controls and communication partner education (Bloch et al., 2011). Communication partner training in supportive communication techniques can increase participation through functional communication and promotion of well-being (Hanson et al., 2011; Simmons-Mackie et al., 2016; NICE, 2023). Communicative participation should be a primary focus of patient-centred intervention (Page & Yorkston, 2022) and should start in the acute hospital. If speech alone is insufficient to meet the individual’s communication needs, a variety of low tech (Garcia et al., 2004; Hustad et al., 2003) and high-tech augmentative strategies should be trialled, subject to the individual’s abilities (Spencer et al., 2006; RSCLT, 2024). The acute rehabilitation hospital setting directly influences the opportunities patients have to practise adaptive strategies and have positive experiences (Clarke & Black, 2005). Staff training can also minimise the negative attitudes patients experience (Fox & Pring, 2005) which can lead to developing isolating, avoidance strategies (Walshe, 2010). Some argue that SLTs include psychological care as an integral part of holistic patient care (Scott & Barton, 2009), and that strategies to address psychosocial impact should be integrated into any program of therapy (Burns, 2009).

### Patient-Centred Approaches and holistic goal setting

The goal of SLT in the acute hospital is to address the multifactorial impact of dysarthria and support the individual to achieve a method of communication appropriate to their medical and social circumstances and priorities (RCSLT, 2024) in preparation for their discharge home. Patients who are actively involved in setting goals and planning their therapy perceive their therapy as more meaningful and effective (Scobbie et al., 2021). However, patients who are only just coming to terms with their disability may feel overwhelmed and not know what they will ultimately find most beneficial. This may go some way to explaining the difficulty demonstrating the impact of early SLT on longer term measures of activity and participation (Mitchell et al., 2017). Social reintegration after stroke involves transitioning through a series of changing goals that may be quite different to goals initially set in the acute hospital (Wood et al., 2010).

To facilitate realistic and achievable goal setting, it is essential that information and education is provided to the person with dysarthria and their significant other(s) about the impact of their dysarthria (MacKenzie et al., 2013) and possible causal and maintaining factors (RCSLT, 2009). Information provided in an accessible format (Stroke Association, 2006), as soon as the patient is medically stable and able to absorb it (Pindus et al., 2018), is reported to reduce anxiety and increase the person’s sense of control and it should be considered key to a successful discharge home (Ellis-Hill et al., 2009).

People with dysarthria describe the usefulness of SLT, of learned techniques and coping strategies, resulting in improved confidence (MacKenzie et al., 2013). However, the differing motivations and barriers to participation reflect the complex reality of participation for people with dysarthria (Scobbie et al., 2021) and may account for the difficulty maintaining gains once SLT finishes (Yorkston et al., 2017). Addressing these should start in the acute hospital phase with the patient, their significant other(s) and staff, by ensuring the intervention is as functional and meaningful as possible (Bright, 2018). Walshe et al. (2009) suggest asking speakers to rate speech relative to other worries and concerns at that time. Different approaches to intervention will have different relevance at each stage in the individual’s recovery (Miller et al., 2017) and indeed delaying intervention may be more beneficial for some (Palmer & Enderby, 2007; Bowen et al., 2012). Others benefit from the early introduction of adaptive strategies, to make sense of their stroke and to preserve their sense of identity (Scott & Barton, 2010), so that they are better able to engage with rehabilitation.

### Context and rationale for research

While previous studies have improved our understanding of the experience of living with dysarthria (Dickson et al., 2008; Walshe & Miller, 2011), there is still limited information about patient perceptions of the speech therapy they have received and its effectiveness, particularly regarding psychosocial factors and its impact on participation.

Hesketh et al. (2011) report that SLTs, patients and families often have different perspectives on what is important in therapy. Therefore, patients have a crucial role in planning and evaluating their own treatment (Roddam & Skeat, 2010) as they are uniquely placed to provide an opinion as to its value (The King’s Fund, 2012). Patient-centred goal setting and service evaluation improves the effectiveness of therapy and the patient’s quality of life (Rautakoski, 2010), with clients who feel that they are heard reporting more effective treatment and greater recovery (Byng et al., 2002).

Bloch et al. (2011) highlight the individuality of experience and suggest caution in assuming that people with the same condition necessarily share common experiences. However, qualitative approaches have been shown to increase phenomenological understanding (Murray & Harrison, 2004) and lessons learned from a detailed exploration of patient experience can be used to improve patient care (Locock et al., 2020).

## AIM

This study aims to explore the participant’s experience of their dysarthria, its impact on their rehabilitation journey and to develop an in depth understanding of their experience of the SLT they received in hospital and how effectively it addressed their needs.

## METHODOLOGY

A phenomenological approach utilizing semi-structured interviews was adopted to better understand the lived experience of patients with dysarthria. An open and non-judgemental phenomenological approach has been argued to effectively bridge the gap between health care practice and research (Finlay, 2011), by focussing on the experience of the patient (Smith et al., 2009). Unlike other forms of qualitative research, such as grounded theory, phenomenology does not attempt to categorize behavior or generate theory. Rather, in focusing on rich, in-depth personal accounts, phenomenology aims to better understand how that person’s world is lived and experienced. This is particularly useful to explore a largely invisible condition (Murray & Harrison, 2004; Carel, 2016), such as dysarthria. Others may perceive minimal impairment, but the person does not feel themselves or is concerned with how they may be perceived by others (Cant, 1997). Phenomenology’s holistic approach provides a framework for understanding the complexity of the experience of dysarthria and the interaction between physical and psychological health (Finlay, 2011). This approach also fits well with the skills and professional values of therapists with a focus on holistic goals, improving communication and enabling people. The relationship of the researcher/interviewer and the service user/participant is acknowledged and recognised as a strength and the service user is seen as a co-collaborator.

While it is recognised that communication impairment may impinge on the data generated, interviews were selected as the data collection method for this study to provide insight into experience as socially and culturally produced (Dickson et al., 2008). Arguably, the phenomenological richness of description comes not only form the words used, but the gestures, tone of voice and other paralinguistic cues which accompany them. Therefore, both verbal and non-verbal data is incorporated into the transcription using interview notes to ensure completeness.

The thematic analysis was framed through a critical realist lens with an inductive approach to develop themes. Critical realism is emancipatory in that it views research as a valid means of improving understanding in order to improve practice (UCL, 2019-Present). This focuses the analysis on the aspects of the interview data most relevant to the research aims (Noble & Smith, 2014).

## METHOD

### Participants

Purposive sampling was used to recruit a range of participants and facilitate comparisons (Dickson et al., 2008). Participants were recruited from the acute hospital SLT caseload with a new diagnosis of stroke, to ensure exploration of their lived experience, unaltered by later reflection. Patients with confusion, delirium or significant language impairment were excluded (score of less than 25/30 on the Frenchay aphasia screen test (Enderby et al., 2013)), to ensure informed consent and the ability to participate fully in the interviews. Age, ethnicity, and previous stroke were not exclusion criteria. Ethical and research and development approval was obtained from the Health and Research Authority. Following screening of the acute caseload, ten patients that fit the criteria were provided with verbal and written information on the study (Appendix B) with 9/10 giving signed, informed consent prior to participating. Nine participants were interviewed between July and October 2024, with the data for one excluded as their diagnosis changed to another neurological condition.

### Data generation and analysis

The semi-structured interview was shared with existing patients prior to finalisation to ensure an easy-to-understand question format (Appendix A). Semi-structured interviews were used to ensure the data gathered was relevant and of sufficient depth and the researcher’s interpersonal skills were employed to keep the focus of the interview on the topic while encouraging open reflection.

Interviews all took place in a quiet room on the acute stroke ward, during their in-patient stay, with the time post stroke to interview ranging from one week to four months. Communication support was provided, including white board and pen, closed questions and reflecting comments back to check and clarify meaning. While this was intended supportively, it is recognised that any misinterpretation could alter the participant’s meaning, a potential methodological weakness. Researcher subjectivity, as an SLT, is acknowledged, but viewed here as a resource in a supportive, interpretive interview style and when analysing data (Gough & Madill, 2012). Synchronistic reflexive journaling was used to ensure ongoing reflexive analysis and acknowledge prior assumptions (Braun & Clarke, 2021).

The data was analysed with Braun and Clarke’s (2021) reflexive thematic analysis to identify themes in the data due to it’s potential for interpretative, theorised analysis (Braun et al., 2022). Consistent with the critical realist approach taken here and qualitative research values (Finlay, 2021), researcher subjectivity was incorporated to develop compelling interpretation grounded in the data (Braun et al., 2022). The analysis continued beyond description and conceptual ordering in order to create learning for implementation in practice (Harding & Whitehead, 2016). The researcher, as a clinician, has an invaluable interpretive role in the attribution of significance to patterns of meaning and the outcomes of the research process (Braun et al., 2022).

## THEMATIC ANALYSIS

The six phases of Braun and Clarke’s (2021) reflexive thematic analysis were used through an iterative process: familiarization with the data, generating codes, constructing themes, revising and defining themes, and writing the report of the analysis.

### Phase 1: Familiarisation with the data

Transcribing the dysarthric speech and incorporating the non-verbal commentary provided much of my initial engagement with the interview data as it required immersion in each interview. Speech irregularities often slowed the transcription process down, with a need to listen multiple times to ensure accuracy. This led me to notice things that hadn’t been apparent when carrying out the interviews and led to greater understanding of the participant’s meaning. Some attempt to include an understanding of their (latent) meaning or relevant or emotive comments was included during initial transcription at the height of understanding as it was felt that distance would not benefit transcription/interpretation. This interpretive element of transcription is recognised in qualitative research (Bird, 2005). Conducting the interviews myself required engaging with participants and building rapport to explore their experience. Therefore arguably, the interpretation started in the interview during reflection of their comments to check and clarify their meaning. Braun et al. (2022) argue that interpretative depth lies in the skill of the analyst, not the method and this seems to be particularly relevant for these participants who often express so much more than their words convey. Reflexive journaling was valuable at this stage to support understanding of the material and insights were added concurrently, to capture my own preconceptions and views as a clinician at the time.

### Phase 2: Generating Codes

Codes were generated in an inductive and iterative process to keep an open mind and learn from the data, rather than using existing theories to guide the interpretation. Coding was generated manually in lieu of computer software, to optimise the intuitive and reflective processes needed to analyse qualitative data (Harding & Whitehead, 2016). Data examples were repeatedly modified to ensure they captured meaning in a way that could be understood in isolation.

### Phase 3: Constructing themes

Themes were constructed from commonalities in the coding and patterns in the data set as a whole. Extra weighting was not given to codes with more examples as each interview was slightly different. Rather themes were constructed where they reflected a shared meaning or experience of the participant group. Excel provided an easy method of sorting and organising the data visually to ensure internal consistency of each theme.

### Phase 4: Developing and reviewing themes

Fourteen initial semantic themes were developed and expanded at this stage to incorporate more latent, interpretative themes. The interview data was returned to regularly during this process, listening to the participant’s own words to ensure their experience was interpreted accurately. This was particularly useful when there was conflicting or overlapping opinions expressed. An interpretive stance was also useful when participants struggled to find words to describe their experience, as it allowed themes to be developed from what participants *were* able to say in association with previous literature (Braun & Clarke, 2006). The thematic analysis is inevitably influenced by the researcher’s clinical experience and theoretical positioning, which required ongoing ethical consideration and reflection.

### Phase 5: Refining, defining and naming themes

Extra time for reflection, provided valuable perspective and new clarity in refining and naming the final themes. Each stage from the initial coding through theme development was recorded to ensure defensibility (Braun & Clarke, 2021). Discussion of the developing analysis with a supervisor experienced in qualitative research was critical to clarify theme descriptions. Articulating the developing themes, enabled the recognition and acceptance of where themes merged and overlapped, avoided confusing topics with themes and avoided premature analytic closure. Participant quotes were chosen to support theme names to ground the themes in the data and give the participants voice.

### Phase 6: Writing the report

The exact nature of the themes continued to be refined during the writing process as reference to relevant literature deepened the understanding of the patients’ experience and the implications for clinical practise. The iterative process of reflexive thematic analysis means that the cyclical reflection and interpretation continued throughout the writing process (Petty et al 2012). Illustrative quotes were selected from the interview data to ensure constructed themes accurately represent the interview data.

## RESULTS & DISCUSSION

The thematic analysis of interviews identified three prominent themes. The themes are multifaceted and therefore inter-related, with each theme adding to the understanding of the participants’ experience: speech as a competing priority in the acute phase, feeling disempowered in rehabilitation, and the need for human connection.

### Theme 1 Speech as a competing priority: “I’m not getting out of hospital if I’ve got perfect speech, but my legs don’t work. You know?”

#### Speech difficulties minimised in the wider impact of stroke

For many, a stroke is a major life event, with a pervasive impact on multiple aspects of functioning which takes time to come to terms with. Participants describe feeling “devasted” (D) and difficulties of “acceptance” (F). Emotionally and cognitively, they may initially struggle to adjust, meaning they simply aren’t ready for SLT:

> *F: the brain has not accepted the fact that this side doesn’t work*.

> *D: I suppose to get my brain back into the habit of, you know? Trying to, trying to do things*.

> *C: They tell me, but I can’t remember*.

Stroke generates a grief response, including negotiation in conceptualising their experience. There is a sense of feeling lucky that they have survived and that their symptoms could have been worse, leading to a tolerance of their dysarthria or a minimisation of its importance. Some only considered their speech to be an issue once they saw SLT, which may account for issues reported elsewhere with motivation for speech rehabilitation (Scobbie et al., 2021):

> *A: You’ve got some speech and there’s that a feeling of, well, at least I can speak once you’ve been told you’ve had a stroke. You think. Well, thank goodness I can still speak*.

> *A: I think that was the first time you kind of thought. Oh, wait a minute. We’re working on my speech here*.

> *B: But then you think. A lot more could have happened. D’you know?*

> *G: I do think I’ll get better. Better. I don’t think I’m bad. I don’t think I’m. I’m sure I’m so much luckier than many people*.

#### Speech as a competing priority

Consistent with previous reports (Olofsson et al., 2005), getting home is a common goal for the participants, which despite their disabilities, represents a freedom and independence that they do not feel in the acute hospital environment. Speech is not perceived as having an immediate causal link with this goal. Dysarthria is only one of several competing priorities in the acute rehabilitation phase and may represent a relatively minor impact on their current function:

> *E: Long as they understand you*.

> *B: I think, it’s not that bad that people would em think that was a point. I think it’s oh, I don’t make her out very well. But I don’t think it’s that bad*.

These accounts are heavily situated within the wider rehabilitation context of the acute hospital phase, with competing needs of physical impairment including mobility and swallowing, continence, language and cognitive recovery factoring strongly in participant interviews. Co-existing conditions such as visual impairment, anxiety, facial weakness and language difficulties compound communication difficulties:

> *D: total weakness on my left side of my face which is, concerning me all the time*.

> *B: I think it’s…the thing about speech. I’m having to try to think of the <u>words</u>. Sometimes the word I want won’t come*.

The challenge of co-existing word finding difficulties associated with language impairment (dysphasia) in addition to reduced speech intelligibility due to dysarthria is prevalent post stroke, with Flowers et al. (2013) reporting co-occurring dysarthria and dysphasia in 15% of their population studied. Participant B’s emphasis on “*words*” illustrates that the preciseness of speech can be secondary to the frustration of not being able to think of the words to convey her meaning.

Participants had differing views of the importance of SLT for dysarthria, depending on the perceived severity of dysarthria and incidence of co-existing conditions. Some reported that they would value daily input, while others did not feel the need for ongoing therapy for what they perceived as minor speech changes. Interestingly, expressed views on frequency of intervention rarely acknowledged limited energy resources, with the same participants who valued daily input noted to have declined offered sessions due to fatigue. SLT sessions themselves were viewed as tiring “to do physically” (B) and they felt “cheated” (A) if they were unable to fully participate. This highlights the importance of good sleep hygiene and scheduling in acute rehabilitation settings (quiet rooms, minimising noisy technology, scheduling therapy sessions to allow protected rest periods), so engagement is not undermined by fatigue:

> *G: I could have cried. I was so tired…You’ve no idea how much I want to [participate in therapy], but I am just so tired*.

> *F: I’m exhausted and I’m doing nothing*.

> *A: You can run out of energy, so you don’t get as much from the session because actually you’re so tired*.

Given patient’s limited energy resources, SLTs must prioritise goals and the initial focus is often on co-existing dysphagia or dysphasia. Rehabilitation has been criticized for its preoccupation with the physical aspects of care (Brady et al., 2016), but participants recognised swallowing as the more pressing medical need and acknowledged the need for a “balance” (E) for therapy sessions. For those whose acute hospital phase is longer, either their physical recovery progresses, or the balance of priorities may shift, and the importance of effective communication emerges as they adapt and cultivate well-being within their illness (Carel, 2016). Once the physical needs are met, the focus may shift to more psychological and social needs (Wade, 2024):

> *B: It was all just too much. And I wanted to keep going [to therapy] more, but my eyes really. I thought, maybe my voice would clear, but it didn’t*.

> *SLT: How important is your swallow? E: Very important. SLT: How important is your speech? E: I want people to understand me*.

> *SLT: So what changed your mind about going to the group? Did you just get bored? A: I think it was something different to do…You couldn’t sit here all the time. You have to progress*.

> *C: I think that where I am at the moment, if I’ve got something specific then I’ll say it, but I think. How could I make things better?…you can’t keep dribbling on*.

This shift in priorities underscores the evolving nature of rehabilitation (Wood et al., 2010), where meaningful patient centred activity fits the needs of the individual at any given time. This may require a combination of approaches, developed in partnership with the patient, which adapts as the patient’s needs evolve:

> *SLT: What do you think was the most effective advice or the most effective approach? A: Every person would be different…I don’t think I’d like to say any one was better than another. I think. You know. Putting it all together*.

#### Limited recall of SLT content

Reduced readiness for therapy and feeling over-whelmed with competing priorities is evidenced by the limited recall participants had of the content and rationale for SLT sessions. When asked what they did with the speech therapist, participants had a vague recall of doing “exercises” (D, E, G & H), being given advice in “some sheets” (G & H) and a goal being to “pronounce words correctly” (E & H):

> *C: Eh, well. I was seeing a speech therapist. Just to try to make me sound clear. SLT: Yeah. How did you do that? C: I don’t know. Not really. I’m sure there were some secrets or so. Anyway. I don’t know. I got something at the time*.

> *SLT: So why do you think therapy is focused on the breath like that then? B: I don’t know, I really. Not technically I don’t, it’s. I can see why breathing is important in all you do. It’s em, kind of ration things out, as it were*.

> *SLT: What do you think has been the most effective therapy or the most effective advice that you’ve had? About speech. D: I’m not sure. Just the whole the whole package has been absolutely wonderful*.

However, this does not appear to represent a lack of value attributed to speech therapy, as there was a consistent appreciation of SLT for dysarthria as being “very helpful” (A, D & F), “absolutely brilliant” (G) and a perceived helpfulness of intervention:

> *B: It gradually got better. I try to speak more slowly now*.

> *C: A simple thing to do [underlining key words to emphasize when reading aloud], but it worked*.

> *H: I change the way I say it…Cos I practise*.

### Theme 2 Feeling disempowered in rehabilitation: “I’m here! You know, I’m not, you know invisible.”

#### A sense of disempowerment

Stroke can result in a profound sense of disempowerment due to its sudden unexpectedness, subsequent disability, and loss of independence. Dysarthria compounds this sense of disempowerment as it impacts the means of self-expression and sense of control over one’s life. Dickson et al.’s (2008) suggestion that the sense of disempowerment manifests both internally through feelings of frustration, negativity and helplessness, and externally, as well-meaning family, friends and healthcare providers, make assumptions or speak on their behalf was supported by the participant’s comments:

> *F: Yeah, a couple of friends have been coming in regularly and you know I don’t have anything to talk about except. It’s all about me. Me, me, me, me. Boring. Boring not. It’s not good. SLT: You don’t like talking about yourself? F: Not when it’s all negative. SLT:*

> *Hmm…And do you feel quite negative at the moment? F: Very. I don’t see much positivity in anything*.

> *G: So you say something and she’ll go “What? What d’you day?” or she’ll make up something that she thinks you’ve said*.

> *E: You know what you’re trying to say, but there’s no-one can*.

#### Healthcare as disempowering

It has been argued (Jones et al., 2000) that health care inadvertently disempowers stroke patients by prioritising physical needs, which may be prerequisite for discharge. In the context of the acute hospital, disempowerment goes beyond self-expression or agency, to institutional disempowerment of the individual. Despite best intentions to ensure patient centred goals, the therapy regimen and daily routine, is not set by the patient, nor do they typically have a say in who sees them and when. Service pressures and time restraints mean that the freedom of the individual as a questioning, autonomous person, can be secondary to the safe and efficient running of the ward or stroke unit (Olofsson et al., 2005). The following comments illustrate that rather than presumed capacity to make their own choices, there is an implicit suggestion of external factors dictating what is in the individual’s best interest and that a “good” (F) patient ought not to question this:

> *A: And say I’m just going to get that to yourself. And the other half says: No, you can’t, you’re not allowed to*.

> *A: I realise it’s for your own good*.

> *F: I guess they’re going to make me do exercises that I don’t want to do.*

> *F: I did well in Physio yesterday and I made no fuss. I was very good*.

Whether necessary or habitual, passive dependence on others to interpret or anticipate one’s needs can result in a feeling of being infantilized. Rather than promoting self-management, this dynamic of competent/professional and incompetent/patient perpetuates a cycle of disempowerment (Hilari & Northcott, 2006), as patients lose confidence in their own ability to communicate choices, become more passive and withdraw from social interaction (Comrie et al., 2001), reinforcing their sense of inadequacy. This is consistent with the prevalence of negative self-talk amongst the participants in this study:

> *A: I feel as if I’m a child again…I mean I need help… It’s very frustrating*.

> *SLT: And what opportunities have you had to practise your speech in the ward? What about like conversational speech with the other patients? H: I’ve not done that. No. I just listen*.

> *SLT: What do you say to yourself? G: Get a grip…So you’re the worst sometimes.*

> *C: I’m no very good am I?*

Self-esteem and identity are vulnerable to the disruption to social interactions caused by dysarthria and negative experiences undermine confidence in communication abilities (Miller et al., 2006). Rehabilitation of speech and communication necessitates a careful balance between supporting and challenging the individual. While verbal encouragement and positive feedback are critical to the rehabilitation process, they are insufficient for instilling a strong self-belief (Hole, 2014). Positive communication experiences are required for this, to resolve the cognitive dissonance created when the patient’s expectations and beliefs about their capabilities don’t match their experience of communication difficulties (Wood et al., 2010). Rehabilitation is typically non-linear, with more challenging experiences temporarily reducing confidence until eventual success builds confidence. Creating positive communication experiences is argued to help preserve self-esteem and identity by supporting the construction of a new, equally valued identity (Wain, 2008):

> *F: I don’t feel like my old self, no*.

> *B: I’ve had to em revise and kind of re-think, things. I’m trying to build a picture of life here as opposed to at home*.

> *SLT: Do you ever just give up? E: No*.

> *C: You know you always try to. Do better*.

> *G: I had to like rein myself in and say it again. And I’d say it again and that was fine*.

#### Importance of hope and positivity

Generally, however, there was a marked expression of optimism and resilience. Within a holistic, patient-centred approach, the importance of psychological gains is recognised (Wain, 2008). The dilemma for therapists, is how to balance the need for positive self-belief with providing education about the condition and managing expectations of recovery. Participants described the importance of positive thinking and hope and were quite accepting of the uncertainty of prognosis. Indeed, when asked what advice they would give to others who had recently experienced a stroke which resulted in dysarthria, they were unanimously positive:

> *A: Everybody should sort eh remember how they were and then suddenly…convince us…by using…these various steps…That it [speech] will come back. But it won’t happen overnight*.

> *E: It [speech] will come back when it wants to come back…You just don’t know when*.

> *G: I felt and I still feel that it [speech] will come back. Yeah, you have to hope…You have to have positivity*.

> *D: It [dysarthria] can be treated*.

This suggests that SLT has a vital role in ensuring the psychological benefits of hope, a sense of agency and in empowering the individual to navigate the acute hospital setting despite their communication challenges. This has important implications for the message given to patients about recovery from dysarthria, as hope of a possible recovery may be an individual’s main agency for motivation (Hole, 2014). Furthermore, group interventions which support connections between patients at an early stage in their dysarthria recovery with those at a later stage may have therapeutic benefits which go beyond practising speech strategies and help address psychological and emotional needs:

> *A: It’s enjoyable to listen to somebody [another patient in group], to see what he’s done with his life and then you realise his life’s been turned upside down. And then to see how well he’s progressed. It gives you wee bit [emotional] more hope and well, there is a way out, you know?*

### Theme 3 The Need for Human Connections: “Sometimes you feel pretty useless, and you have nothing to talk about. Which somehow makes you less of a person.”

#### The need for human connections

Speech difficulties during the acute hospital phase can result in a cycle of reduced opportunities and reduced confidence for social interaction and conversation. The fundamental need for human connection is reflected in the participants comments and how much they value visits from friends and family:

> *F: I know I would have gone clean round the bend, if my daughter hadn’t come in every day. And we don’t chat for long*.

> *B: I just like to see as many people as want to and can come. You know. I know two of my friends can’t come now. They’re not able and I really feel quite sorry about that*.

Even limited social interaction has a clear impact on the sense of one’s place in the world and can help to maintain friendships and re-establish social identity. When speaking over the phone is too challenging, text messaging is a valued substitute and when additional visual or language difficulties preclude this, exchanging photos can be enough to show they’re thinking of them and maintain relationships:

> *B: Even if it’s eh “Hello”. I’ll know it’s you and eh, say I’ll fine or…or something. And that’s enough. I don’t have long conversations*.

> *F: We have photos um messages back and forth*.

Extra weight is put on feedback received from friends and family regarding their speech clarity, often surpassing their own judgement. Indeed, there is the perception that so long as they are understood by the people that matter, there is a tolerance for altered speech:

> *A: Or [family] maybe just said your speech isnae bad*.

> *SLT: So do you listen to [husband] when he tells you how your speech is. H: Yes, more so. G: As long as they’re understanding me. It’s not a priority, fine. They’re understanding [me]*.

> *G: Nobody in the hospital knows me. Em. But all my family and friends know what I sounded like before and what I sounded like now. So it was their feedback I’m waiting on. Em and I have told them all, I’m wanting their feedback. I want their honest feedback*.

Communication effectiveness may depend on who they’re speaking to and the ability of significant others to “tune in” (B) to speech acts as a protective factor. While some report communication with family to be “very difficult” (H), close family may use an element of anticipation to support understanding, even if they only get it right “sometimes” (H):

> *A: Or if they don’t [understand] and I’ve got to repeat things. I think I’ll be concerned then*.

> *B: I don’t know. [Son] was saying this morning, he thought my speech wasn’t bad at all. But he’d say that. It could be he’s just well tuned in*.

> *H: They [family] can communicate. They know what I’m going to say there*.

Out-with the immediate support network, concern about not being understood or being perceived as less intelligent can lead to negative strategies, a more passive role in conversation or avoiding conversation altogether. Speech may be objectively quite intelligible, but this doesn’t appear to encourage social participation if it “doesn’t feel like it” (B):

> *C: No, they’ll think daft me.*

> *B: I’d rather listen really*.

> *A: I suppose I’ll…I’ll be right wary. If they’re understanding me*.

#### SLT as surrogate significant other

Successful rehabilitation requires supporting positive participation in meaningful, functional situations. While 1:1 conversation practise was perceived as “very helpful” (A), and “definitely needed” (B), the benefits of SLT appear to go beyond this, with the SLT at times becoming a surrogate significant other. This is valuable to the rehabilitation process as an empathetic therapeutic relationship can enhance motivation (Wain et al., 2008) and provides a safe and supportive environment to practise communication strategies and build trust in the rehabilitation process (Bright, 2022):

> *B: Oh I definitely needed that [1:1 support].*

> *E: Make you feel more at ease*.

> *A: And you can get through to me a lot easier.*

> *H: She [SLT] can pull you up harder*.

#### Therapeutic value of wider rehabilitation setting

Subjective beliefs about the potential for recovery are integral to the rehabilitation process (Hole, 2014). Negative experiences undermine hope and motivation (Burns et al., 2017) and should be minimised by a multi-disciplinary team with a good understanding of needs, practical solutions, and supportive communication skills. The value of interaction with a good communicator is recognised (Young et al., 2012), and positive experiences of being understood despite dysarthric speech can reinforce the importance of communication effectiveness as opposed to speech clarity:

> *A: Well, I think this wee building that you’ve [emotional]. Is a god send to [emotional] to people like me*.

> *G: As for the speech. I think it’s getting. I think it could be a lot worse. Em. And I just need to get on. Keep practising*.

> *F: I just got a message while [the nurse] was here and I said “Ohh dear I can’t read that, it’s too small”, and she said let me sort it for you and click click, click and she did it*.

> *D: And and my mouths not the only part of the body, you know, so I can speak with my hands to to really*.

> *SLT: What was interesting was when we did that test, looking at the words. Where you had to name all the different pictures. And you were brilliant. E: I knew they [the words] were there*.

This latter quote came from a participant who spoke very little initially, due to a severe dysarthria rather than a language difficulty, but turned a rehabilitation corner after hearing himself successfully naming pictures in a book.

#### Re-constructing identity and social role

Enabling positive social experiences not only helps with self-esteem but allows the person to start to establish who they have become and where they may now fit in their social sphere:

> *B: Somebody said to me this morning that I was quite patient and I hadn’t realized that part of me. I didn’t look on myself as a patient person. But they must find that*.

However, participants report feeling less able to participate in social conversation because they feel they have nothing to talk about except their illness:

> *F: Yeah, a couple of friends have been coming in regularly and you know I don’t have anything to talk about except. It’s all about me. Me, me, me, me. Boring. Boring not. It’s not good*.

A stimulating environment is key to rehabilitation (Roding et al., 2003). Therapy and activity groups provide an opportunity to interact in a safe space, practise strategies and be with others “in the same boat” as them. Peer support in groups can improve self-esteem and motivation for recovery (Palmer & Enderby, 2007) by supporting reflection on their impairments and recovery journey:

> *B: Well if I thought that em people [in the group] weren’t understanding what I’m saying, I think I would try even harder. I wouldn’t give up*.

> *G: And you’re thinking, just for that second, there’s that doubt in your mind that you’ve said something different…That you’ve said something that she didn’t understand…Wee devil sitting on your shoulder. Whereas now? I’ve realised other people are talking to her and the same thing’s happening. It’s not me at all!*

> *C: I could sit back. Or I could come across with something.*

> *A: It’s a better environment. It’s a more normal environment*.

> *D: It helped me to realise that I’m not the only person in… the… world… that has this problem. It’s a recognised thing with the stroke*.

Participants describe a deep desire to re-establish and maintain social connections, finding comfort and hope in positive interactions with family, friends, and peer support networks. Visits from friends and family are crucial for motivation and resilience (Northcott & Hilari, 2013) and quality interactions with healthcare staff and other patients should be valued and encouraged.

## CONCLUSION

This study illustrates how the experience of dysarthria and dysarthria therapy cannot be isolated from the multidimensional nature of stroke rehabilitation generally, which touches upon the interrelated physical, emotional, and social dimensions of patients’ lives. The researcher’s clinical role as an SLT means these findings inevitably result in critical reflection on practise, and the implications for SLT are considered here.

Despite the crucial role of communication in social reintegration and the consequences of dysarthria for emotional and psychological well-being, patients may perceive speech difficulties as a secondary issue in the acute phase, often demonstrating a degree of acceptance of their altered speech. Limited patient and therapist resources means priority may be given to goals which are perceived as more closely related to the goal of getting home/getting back to normal. However, patient priorities evolve over the rehabilitation journey and SLT intervention must adapt alongside to maintain motivation, including addressing social and emotional needs.

Communication difficulty results in loss of autonomy, self-esteem, and disrupts identity/social role making it difficult to navigate the rehabilitation context independently. This results in a cycle of disempowerment in the acute hospital setting which fails to adequately prepare the patient for their discharge home. To counteract this, acute hospital rehabilitation must seek to empower the patient throughout their rehabilitation journey, with all healthcare staff being aware of beneficial and potentially harmful forms of interaction. A holistic, patient-centred approach to rehabilitation should explicitly address the psychosocial elements of stroke rehabilitation with positive social experiences. Failing to incorporate meaningful activities to practise communication strategies, build self-esteem and promote self-management could undermine attempts at rehabilitation.

The expressed need for human connection further emphasises the importance of social activities in rehabilitation, both as a means of practising communication strategies and as a means of building self-esteem and motivation. Ongoing service pressures encourages innovation, and the embracing of electronic therapy delivery and telehealth may seem a potential solution. However, this study suggests that rather than attempting to optimise therapy dosage with electronic devices, skilled multi-disciplinary teams, patient groups, and significant others from their social support network should be integrated into therapy plans to reconstruct a social identity and establish self-management strategies for communication prior to discharge.

Patient-centred dysarthria therapy in the acute hospital phase has a vital role in helping stroke survivors adjust to new communication difficulties and regain confidence and independence. However, challenges related to competing rehabilitation demands, the psychological impact of stroke and the complexity of patients’ conditions necessitate a compassionate, holistic, and flexible approach to SLT intervention. Future research should examine how holistic and collaborative goal setting can be effectively implemented in the acute hospital setting and the potential benefits of integrating psychological support strategies with speech therapy for longer term emotional and social participation outcomes.

## Data Availability

All data produced in the present study are available upon reasonable request to the authors

## Appendix A: Semi-structured interview

### SEMI-STRUCTURED INTERVIEW

Hello. Thank you for coming and giving me your time today. We’re going to chat about your speech and about your experience of the speech therapy you’ve received. Can I just confirm that you’re happy for me to record our conversation today? [start recording]

OK, I’ve got four topics I’d like to cover [list topics]. I’ve also got this sheet here to make sure I don’t forget to ask any of my questions. If there’s any question you’d prefer not to answer, that’s fine. We can stop or take a break whenever you like. I just want to learn as much from your experience as possible.

#### EXPERIENCE OF SPEECH FOLLOWING A STROKE

Let’s start at the beginning…when did you have your stroke? (Confirm days/weeks)

Everyone’s experience of a stroke is different, can you tell me a little about how yours has affected you? /How do you think your life has changed since then?

[To redirect] Could we talk a little more about your speech? When did you realise that your speech had changed?

Did you have to adapt to communicate with people initially?

#### EXPERIENCE OF THERAPY

Now let’s talk therapy…how often have you had speech therapy?

Was it the same therapist each time? Did you also see the SLTA?

How long were the therapy sessions?

How would you describe your therapy sessions? What did you work on?

Tell me a bit about that, was that helpful?

Why do you think therapy focussed on that?

What else did you work on?

And what about that, did you find that helpful?

Why do you think therapy focussed on that?

Did you do anything else in the sessions?

And did you find that helpful?

Why do you think therapy focussed on that?

That’s great, you’re giving me lots of wonderful information to work with. I’ve a few more questions and then we’ll be finished.

#### REFLECTING ON EXPERIENCE

When you think back to the first few days following the stroke, how did you feel about your speech?

And how do you feel now?

Is there anything you think you have learnt since then, that you wish you’d known at that time?

(Do you think you’ve had to develop new strategies to help you communicate with people?)

#### REFLECTING ON THERAPY

What do you think was the most effective therapy/advice/approach?

What could’ve been better?

Was there something that you didn’t find helpful or that you didn’t like?

Thank you so much for your time today, it’s been so interesting to speak to you. This will hopefully be useful not just to complete this project, but also to ensure our speech therapy service is as effective as possible.

Would you like me to send you a summary of our findings when the projects finished?

[stop recording]

## Appendix B: Participant Information Sheet

### PROJECT: Patient experience of dysarthria therapy

My name is Sasha Archer and I work as a Speech and language therapist with NHS Lothian.

In order to improve the speech and language therapy service, I am exploring the experience of patients receiving intervention for speech difficulties following a stroke (dysarthria) in the acute hospital phase.

I am completing this research as a student as part of a post-graduate Master of research in speech & language therapy from the school of social sciences at Queen Margaret University in Edinburgh.

### What is the research about?

We want to better understand the experience of therapy for speech difficulties (dysarthria) following a stroke. This will help us to better support others with dysarthria in the future.

### Why am I being asked to participate?

You are being invited to participate in the study, because you have recently had a stroke that has affected your speech (dysarthria) and have had speech therapy for this.

I am looking for 6-8 volunteers to speak to me about their experience. I have a few questions and will give you as much time to describe your experience as you wish.

### Do I have to take part?

No. Your decision to participate or not, will not affect the therapy you receive.

You can take some time to decide or talk to others about the study.

You will be asked to sign to consent to taking part. If you can’t sign, you can make a mark. We will have another person witness this.

### Will I be able to take part?

We want to include as wide a range of experiences as possible, so you can take part even if you find it very difficult to speak or find it difficult to think of the right word. As long as you can understand a few simple questions, we can support you to be heard.

### What are the risks and benefits?

Talking about your experience may trigger some difficult emotions. We will chat in a quiet, private room in the hospital and anything that comes up can be talked through.

It can also be useful to talk about your experience. If you would like to talk to someone in more depth about your feelings, you can also talk to your stroke consultant.

### What will happen if I take part?

I have a few questions to ask. The length of the interview depends on how much you have to say. It may only be 10 minutes, but will not be longer than 60 minutes. The interview will be audio recorded and I will take notes. You can stop at any time and don’t have to give a reason.

You can also chat with me a couple of times (no more than 3 times). It’s your choice. If you wish to speak to me more than once, but leave hospital before we speak again, you will be offered a visit to your home within two weeks to finish our discussion. However, you do not need to continue with the interview if you do not wish to.

### What happens after?

Our conversation will be audio recorded and kept confidential while I transcribe it. The recording will then be deleted. If you choose to withdraw and don’t want your comments used, the recording will be deleted, the electronic files deleted and the notes shredded.

I will need to use information from your medical records for this research project. This information will include:

- your NHS number
- your name
- your contact details
- your medical history

I will only use the information that I need to do the research or to check your records to make sure that the research is being done properly.

Your data will be stored with a code number, so that I can send you a summary of the project’s findings if you wish.

I will keep all information about you safe and secure. I will also follow all privacy rules. All personal data will be deleted within six months.

Once I have finished the study, I will keep some of the data so I can check the results. I will write my report in a way that no-one can work out that you took part in the study. All research data will be deleted within two years.

### What are your choices about how your information is used?

You can stop being part of the study at any time, without giving a reason. If you are happy for me to continue to use your data and comments, I will keep information about you that I already have.

I need to manage your records in specific ways for the research to be reliable. This means that I won’t be able to let you see or change the data I hold about you.

You can find out more about how I use your information:

- at www.hra.nhs.uk/information-about-patients/
- by asking one of the Speech therapy team
- by ringing me, Sasha Archer (Speech Therapy) on 0131 537 3320
- by emailing the QMU Data Protection Officer - The project is sponsored by Queen Margaret University. If you would like to contact an independent person, who knows about this project but is not involved in it, you are welcome to contact the programmer leaders:

Name of adviser: Dr Rosie Arthur and Dr Rachael Davis

Address: Masters of research programme leaders Social sciences division Queen Margaret University, Edinburgh EH21 6UU

Email / Telephone: / 0131 474 0000

If there are any safeguarding or malpractice concerns/an individual is considered at risk of harm, confidentiality may have to be broken in accordance with the NHS Lothian Adult Support and Protection Procedure (March 2023).

If you wish to report a concern or complaint about the study or your experience with the NHS, you can contact:

NHS Lothian Patient Experience Team

Waverley Gate

2-4 Waterloo Place

Edinburgh

EH1 3EG

Tel: 0131 536 3370 or

**Thank you for taking the time to find out more about the project.**

